# Examining the Genetic Links between Clusters of Immune-mediated Diseases and Psychiatric Disorders

**DOI:** 10.1101/2024.07.18.24310651

**Authors:** Sophie Breunig, Younga Heather Lee, Elizabeth W. Karlson, Arjun Krishnan, Jeremy M. Lawrence, Lukas S. Schaffer, Andrew D. Grotzinger

**Author notes:** Corresponding Author: Sophie Breunig, Institute for Behavioral Genetics, University of Colorado at Boulder, 1480 30^th^ Street, Boulder, CO 80303.

## Abstract

**Importance:** Autoimmune and autoinflammatory diseases have been linked to psychiatric disorders in the phenotypic and genetic literature. However, a comprehensive model that investigates the association between a broad range of psychiatric disorders and immune-mediated disease in a multivariate framework is lacking.

**Objective:** This study aims to establish a factor structure based on the genetic correlations of immune-mediated diseases and investigate their genetic relationships with clusters of psychiatric disorders.

**Design, Setting, and Participants:** We utilized Genomic Structural Equation Modeling (Genomic SEM) to establish a factor structure of 11 immune-mediated diseases. Genetic correlations between these immune factors were examined with five established factors across 13 psychiatric disorders representing compulsive, schizophrenia/bipolar, neurodevelopmental, internalizing, and substance use disorders. We included GWAS summary statistics of individuals of European ancestry with sample sizes from 1,223 cases for Addison’s disease to 170,756 cases for major depressive disorder.

**Main Outcomes and Measures:** Genetic correlations between psychiatric and immune-mediated disease factors and traits to determine genetic overlap. We develop and validate a new heterogeneity metric, *Q_Factor_*, that quantifies the degree to which factor correlations are driven by more specific pairwise associations. We also estimate residual genetic correlations between pairs of psychiatric disorders and immune-mediated diseases.

**Results:** A four-factor model of immune-mediated diseases fit the data well and described a continuum from autoimmune to autoinflammatory diseases. The four factors reflected autoimmune, celiac, mixed pattern, and autoinflammatory diseases. Analyses revealed seven significant factor correlations between the immune and psychiatric factors, including autoimmune and mixed pattern diseases with the internalizing and substance use factors, and autoinflammatory diseases with the compulsive, schizophrenia/bipolar, and internalizing factors. Additionally, we find evidence of divergence in associations within factors as indicated by *Q_Factor_*. This is further supported by 14 significant residual genetic correlations between individual psychiatric disorders and immune-mediated diseases.

**Conclusion and Relevance:** Our results revealed genetic links between clusters of immune-mediated diseases and psychiatric disorders. Current analyses indicate that previously described relationships between specific psychiatric disorders and immune-mediated diseases often capture broader pathways of risk sharing indexed by our genomic factors, yet are more specific than a general association across all psychiatric disorders and immune-mediated diseases.

**Key points:** *Question:* How do immune-mediated diseases cluster together and how are they associated with clusters of psychiatric disorders?

*Findings:* Immune-mediated diseases cluster into four factors along the overlapping spectrum of autoimmune to autoinflammatory diseases. There are seven significant factor correlations between immune-mediated disease factors and psychiatric disorders factors along with more specific pairwise associations between two disease traits.

*Meaning:* Immune-mediated diseases are genetically associated with psychiatric disorders. While some associations seem to be driven by more general pathways, we also find that some of the shared signal is more specific to individual psychiatric-immune disease pairs.

## Introduction

Immune-mediated diseases have been phenotypically linked to a range of psychiatric disorders, including eating^1^, mood^2^, psychotic^3,4^, stress-related^5^, and neurodevelopmental disorders^6^. Recent genomic studies indicate that these phenotypic associations are mirrored at the genetic level. For example, an investigation of five psychiatric disorders and seven immune-mediated diseases found evidence of shared genetic signal for 24 out of 35 psychiatric-immune disorder pairs^7^. In addition, the human leukocyte antigen (*HLA*) region of the genome in the major histocompatibility complex (MHC) is critical for the adaptive immune response system and is one of the most implicated regions in psychiatric genetics^8^. Widespread associations across psychiatric and immune-mediated outcomes suggests that overlap identified for pairs of psychiatric-immune disorders will often reflect shared etiology across a broader multivariate system of disorders. At the same time, meaningful biological distinctions within immune-mediated diseases caution against prematurely concluding that all immune-mediated diseases have equivalent associations with psychiatric traits^9^.

Immune-mediated disease encompasses an overlapping continuum from autoimmune to autoinflammatory diseases^9^. These two ends of the continuum share the property that they result from a malfunction of the immune system that includes targeting self-tissue without an observable trigger, such as infection or injury, and subsequent systemic inflammation^10^. They are distinguished by which part of the immune system is malfunctioning: while autoimmune diseases are classified by an abnormal response primarily of the *adaptive* immune system, autoinflammatory diseases are characterized by a malfunctioning *innate* immune system^11,12^. Though they work closely together, the adaptive and innate immune system also diverge with respect to the different types of cells and cell responses that form a complicated network of interactions. Specifically, the adaptive immune system involves T lymphocyte subsets, B cells, and plasma cells, whereas the innate immune system encompasses cells such as macrophages, natural killer cells, and dendritic cells^13^. These divergent biological pathways suggest that autoimmune and autoinflammatory diseases may have similarly distinct associations with psychiatric disorders, which remains to be investigated in a systematic fashion.

Pervasive pairwise associations identified across immune-mediated and psychiatric diseases necessitate a comprehensive multivariate approach to better characterize the shared etiological landscape across psychiatric and immune disorders. Genomic structural equation modeling (Genomic SEM)^14^ is a recently introduced multivariate method that uses the shared features of genomic data to bring together even mutually exclusive datasets into a single statistical model and parse convergent and divergent genetic signal. In this study we apply Genomic SEM to *i*) establish a genomic factor structure of immune-mediated diseases and *ii*) investigate the degree to which these genomic factors of immune-mediated diseases are related to genomic factors of psychiatric disorders. The current study thereby clarifies the relationships between individual immune-mediated diseases and psychiatric disorders by investigating patterns of risk sharing between these two frequently linked classes of diseases on a factor and individual trait level.

## Methods

### Phenotype Selection and Quality Control

The list of immune-mediated diseases from the Mount Sinai Website (https://www.mountsinai.org/health-library/diseases-conditions/autoimmune-disorders), as well as recent studies in the immune-disease field, were referenced to make an initial list of disease traits that was cross-referenced with the CDC website (https://www.cdc.gov/index.html) to verify their status as an immune-mediated disease. This list was further restricted to immune-mediated diseases that were an immediate result of the immune system, rather than a downstream effect of immune activation associated with a different trigger (e.g., eczema). For each phenotype, we selected the most well-powered, publicly available GWAS of unrelated individuals of European genetic ancestry. We removed any traits with existing genome-wide association study (GWAS) summary statistics with low genomic coverage (< 850,000 SNPs) or insufficient power, reflected by a SNP-based heritability 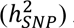 *Z*-statistic < 4^15^. This resulted in a final list of 11 immune-mediated diseases (**Table 1**). For psychiatric traits, the same GWAS summary statistics of 13 disorders were used from a recently described psychiatric genomic model^16^. All GWAS summary statistics underwent a standardized quality control process using the *munge* function within the *GenomicSEM* R package. This function restricts the data to HapMap3 SNPs, aligns all GWAS summary statistics to the same reference allele, and filters at a minor allele frequency > 1% and an imputation quality > 0.9, when available.

**Table 1.** Sample Characteristics of Included Traits.

| Phenotype | Cases | Controls | Liability Scale Heritability |
| --- | --- | --- | --- |
| *Immune Mediated Diseases* |  |  |  |
| Addison's disease (AD)<sup>36</sup> | 1,223 | 4,097 | 16% |
| Celiac disease (CEL)<sup>24</sup> | 2,364 | 324,074 | 13% |
| Crohn's disease (CD)<sup>37</sup> | 12,194 | 28,072 | 22% |
| Diabetes type 1 (T1D)<sup>38</sup> | 18,942 | 501,638 | 14% |
| Juvenile idiopathic arthritis (JIA)<sup>39</sup> | 3,305 | 9,196 | 13% |
| Myasthenia gravis (MG)<sup>40</sup> | 1,873 | 36,370 | 11% |
| Psoriasis (PSO)<sup>41</sup> | 15,967 | 28,194 | 15% |
| Psoriatic arthritis (PSA)<sup>42</sup> | 5,065 | 21,286 | 13% |
| Rheumatoid arthritis (RA)<sup>43</sup> | 14,361 | 43,923 | 9% |
| Systemic lupus erythematosus (SLE)<sup>44</sup> | 4,036 | 6,959 | 10% |
| Ulcerative colitis (UC)<sup>37</sup> | 12,366 | 33,609 | 15% |
| *Psychiatric Disorders* |  |  |  |
| Alcohol use disorder (ALCH)<sup>45</sup> | 8,485 | 20,272 | 14% |
| Anorexia nervosa (AN)<sup>46</sup> | 16,992 | 55,525 | 16% |
| Anxiety disorder (ANX)<sup>47</sup> | 31,977 | 82,114 | 23% |
| Attention-deficit/hyperactivity disorder (ADHD)<sup>48</sup> | 38,691 | 186,843 | 18% |
| Autism spectrum disorder (ASD)<sup>49</sup> | 18,381 | 27,969 | 12% |
| Bipolar disorder (BIP)<sup>50</sup> | 41,917 | 371,549 | 19% |
| Cannabis use disorder (CUD)<sup>51</sup> | 14,808 | 343,726 | 19% |
| Major depressive disorder (MDD)<sup>52</sup> | 170,756 | 329,443 | 9% |
| Obsessive compulsive disorder (OCD)<sup>53</sup> | 2,688 | 7,037 | 30% |
| Opioid use disorder (OUD)<sup>54</sup> | 3,211 | 11,589 | 19% |
| Post traumatic stress disorder (PTSD)<sup>55</sup> | 2,424 | 7,113 | 24% |
| Schizophrenia (SCZ)<sup>56</sup> | 53,386 | 77,258 | 22% |
| Tourette's syndrome (TS)<sup>57</sup> | 4,819 | 9,488 | 22% |

*Note.* This table describes all GWAS summary statistics of all external traits included in the analyses.

### LD-score Regression

Multivariable Linkage Disequilibrium (LD)-score regression (LDSC)^15^ was performed using LD scores calculated from the European subset of the 1000 Genomes Phase 3 project. The MHC region was excluded due to complex LD structures in this region that can bias results. Multivariable LDSC produces two matrices as output: (*i*) the genetic covariance matrix with the SNP-based heritabilities and genetic covariances on the diagonal and off-diagonal, respectively, and (*ii*) a sampling covariance matrix with the squared standard errors of the estimates on the diagonal and the sampling dependencies on the off-diagonal representing participant sample overlap. Since all included traits were binary, the LDSC estimates were converted to a more interpretable liability scale, accounting for the continuous distribution of genetic risk (liability) and sample ascertainment. The liability scale conversion was calculated using the population prevalence specified in the corresponding GWAS paper or a prevalence reflective of the participant sample. Additionally, we used the sum of effective sample sizes across the contributing cohorts for the ascertainment correction, to produce the most accurate estimates of liability-scale heritability^17^.

### Genomic SEM

Genomic SEM takes as input the output from multivariable LDSC along with a user specified SEM^14^. We first modeled the genetic overlap across only immune-mediated diseases. Good model fit, which indexes how well the specified model captures the data, was established based on the comparative fit index (CFI ≥ .9) and the standardized root mean squared residual (SRMR ≤ .1). Our initial model reflected a theoretically motivated two-factor confirmatory factor analysis (CFA), with the two factors defined by either autoimmune or autoinflammatory diseases, with mixed presentation diseases being specified to cross-load on both factors. The two-factor solution had poor model fit (Table S1). We therefore ran a data-driven combination of exploratory analyses (EFA) followed by CFAs. The number of factors to include in the EFA for the immune-mediated disease phenotypes was determined based on the Kaiser rule^18^, the acceleration factor, and the optimal coordinates^19^. To avoid model overfitting, we performed EFA and CFA on the odd and even chromosomes, respectively. Having finalized a four-factor model for immune-mediated diseases based on this approach, this model was integrated with a previously established five-factor model for 13 psychiatric disorders^16^. The psychiatric factors in this model can be approximately described as reflecting compulsive, schizophrenia/bipolar, neurodevelopmental, internalizing, and substance use disorders. The five psychiatric factors were specified to correlate with all immune-mediated disease factors and these inter-factor correlations were evaluated for significance.

We also examined associations between specific pairwise combinations of psychiatric and immune diseases that were not captured by the factor model. This was achieved by adding residual genetic correlations between a psychiatric-immune disease pair to the model. Which residual correlations to add was determined by first inspecting a matrix of model misfit, calculated as the difference between the model implied and observed covariance matrices. This matrix of misfit was then divided by the standard errors on those estimates to create a matrix of *Z-*statistics. Finally, residual genetic correlations between a psychiatric and immune disease were added to the model when there was a *Z*-statistic > 2 for that cell of the matrix. Reported residual genetic correlations are standardized relative to the total SNP-based heritability (i.e., they are not partial genetic correlations). Across all analyses, significance was determined after applying FDR multiple testing correction implemented using the *p.adjust* function in the *stats* R package.

### Q_Factor_ Heterogeneity Test

As part of the current manuscript, we introduce and validate a heterogeneity metric that we term *Q_Factor_.* This metric is a 𝜒^2^ distributed measure of local model misfit that captures inter-factor correlations that fail to recapture the pairwise genetic relationships between the different disorders that define these factors. This could occur, for example, if two traits that define the same factor have directionally opposing relationships with the traits that define a second factor. *Q_Factor_* is specific to a pair of factors in the model, such that multiple *Q_Factor_* metrics can be calculated for a multifactorial model. This new addition to Genomic SEM was validated via simulations that are detailed in the **Online Supplement** and the mechanisms through which significant *Q_Factor_* results can occur are illustrated in Figure S1. For our empirical results, a separate *Q_Factor_* was calculated for each psychiatric with immune-mediated disease factor correlation. *Q_Factor_* was used as a quality control check, where psychiatric-factor correlations are only reported as significant below when *Q_Factor_* was not significant. This is because a non-significant *Q_Factor_* result indicates the factor correlation appropriately describes broad pathways of risk sharing across these two clusters of disease traits.

## Results

### Factor Structure for Immune-Mediated Diseases

The acceleration factor indicated 1 factor should be retained, the optimal coordinates rule 2 factors, and the Kaiser rule 4 factors. While a 1- and 2-factor model did not fit the data well (Table S1), a four-factor model of the immune-mediated diseases fit the data well in all autosomes (CFI = 0.96; SRMR = 0.08; **Figure 1**; Table S1). The four factors can be classified as reflecting an autoimmune disease factor, a single indicator celiac disease factor, a mixed pattern disease factor, and an autoinflammatory disease factor. These factors reassuringly fall onto the theoretically overlapping spectrum described in the literature ranging from autoimmune to autoinflammatory diseases.

**Figure 1.**
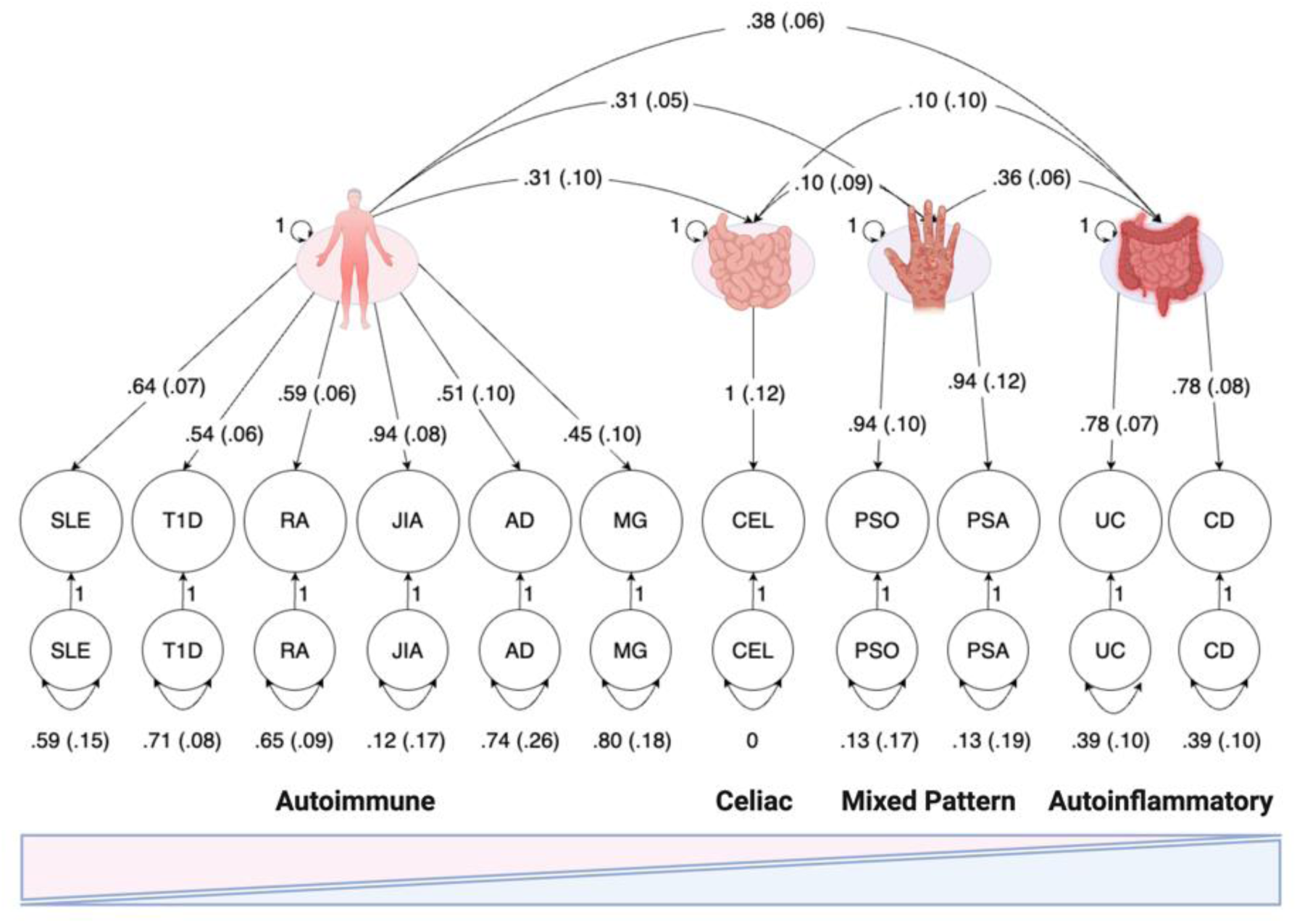
Immune-mediated Disease Correlated Factors Model. Path diagram with standardized estimates for the correlated factors model used in genomic structural equation modelling (genomic SEM) with four factors of immune-mediated diseases. Latent variables are represented as circles. The genetic component of each phenotype is represented with a circle because the genetic component is a latent variable that is not directly measured but is inferred using linkage disequilibrium score regression (LDSC). Single-headed arrows are factor loadings; double-headed arrows connecting back to the same origin are variances; and double-headed arrows connecting two variables are correlations. Paths labeled 1 are fixed to 1; all estimates are given next to the path with standard errors in parentheses. Disease abbreviations are defined in Table 1.

### Genetic Associations Between Immune-Mediated Diseases and Psychiatric Disorders

A factor model that integrated the psychiatric and immune-mediated diseases initially yielded model fit below the recommended threshold for CFI (CFI = 0.88; SRMR = 0.09; Table S1). Significant *Q_Factor_* results for pairs of factors, which we consider in more detail below, indicated that part of this model misfit stemmed from inter-factor correlations that did not fit the data. Residual genetic correlations between pairs of psychiatric and immune disease traits were then added to model where indicated. This updated model fit the data well (CFI = 0.92; SRMR = 0.08; Table S2 for full model output, **Figure 2** for significant residual correlations). Results are described separately below for each immune-mediated disease factor. *Q_Factor_* results are given from the initial model, and significant inter-factor correlations, as well as residual correlations between pairs of disorders, are provided from the final, well-fitting model. We do not discuss the celiac disease factor below as it was not significantly genetically correlated with any of the psychiatric factors, though it was found to be *Q_Factor_*significant with the neurodevelopmental disorders factor (*p_FDR_*= 2.94e-02).

**Figure 2.**
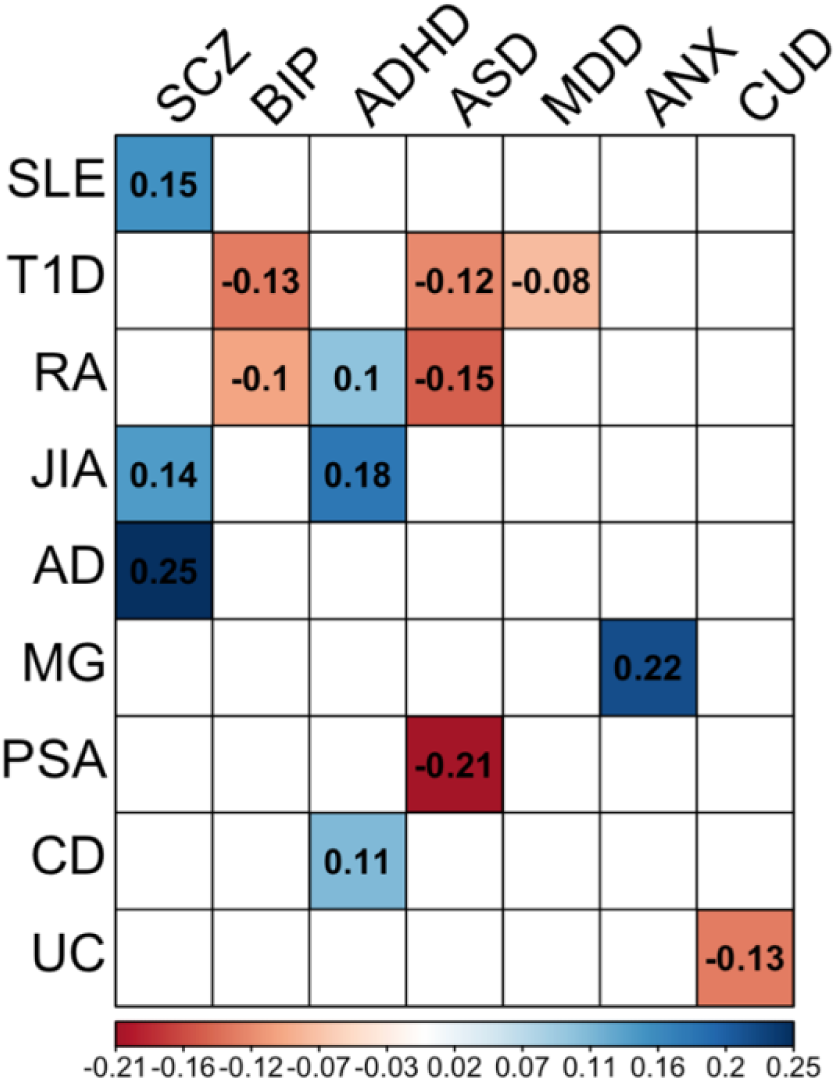
Significant Residual Genetic Correlations. Significant residual genetic correlations between immune-mediated diseases and psychiatric disorders. Disease abbreviations are defined in Table 1.

### Autoimmune Diseases Correlate with Internalizing and Substance Use Disorders

Autoimmune diseases were positively genetically correlated with the internalizing (*r_g_* = .10 [SE = .04], *p_FDR_*= 1.73e-02) and substance use disorders factors (*r_g_* = .12 [.05], *p_FDR_* = 4.38e-02; **Figure 3**). *Q_Factor_* analysis further revealed that there was divergent signal in terms of the genetic correlations between the individual indicators loading onto the autoimmune disease factor and the schizophrenia/bipolar (*p_FDR_* = 2.45e-11) and neurodevelopmental disorders factors (*p_FDR_* = 4.88e-06). The addition of 11 significant residual genetic correlations to the model clarified that these *Q_Factor_*findings were driven by the fact that the disorders that defined the schizophrenia/bipolar and neurodevelopmental factors often showed divergent associations with the autoimmune diseases (**Figure 2**). For example, while schizophrenia showed positive residual genetic correlations with Addison’s disease (*r_g_resid_* = .25 [.05], *p_FDR_* = 6.99e-05), systemic lupus (*r_g_resid_* = .15 [.04], *p_FDR_* = 7.11e-03), and juvenile arthritis (*r_g_resid_* = .14 [.05], *p_FDR_*= 1.73e-02), bipolar disorder was negatively correlated with type I diabetes (*r_g_resid_* = -.13 [.03], *p_FDR_* = 9.49e-05) and rheumatoid arthritis (*r_g_resid_* = -.11 [.04], *p_FDR_* = 1.39e-02).

**Figure 3.**
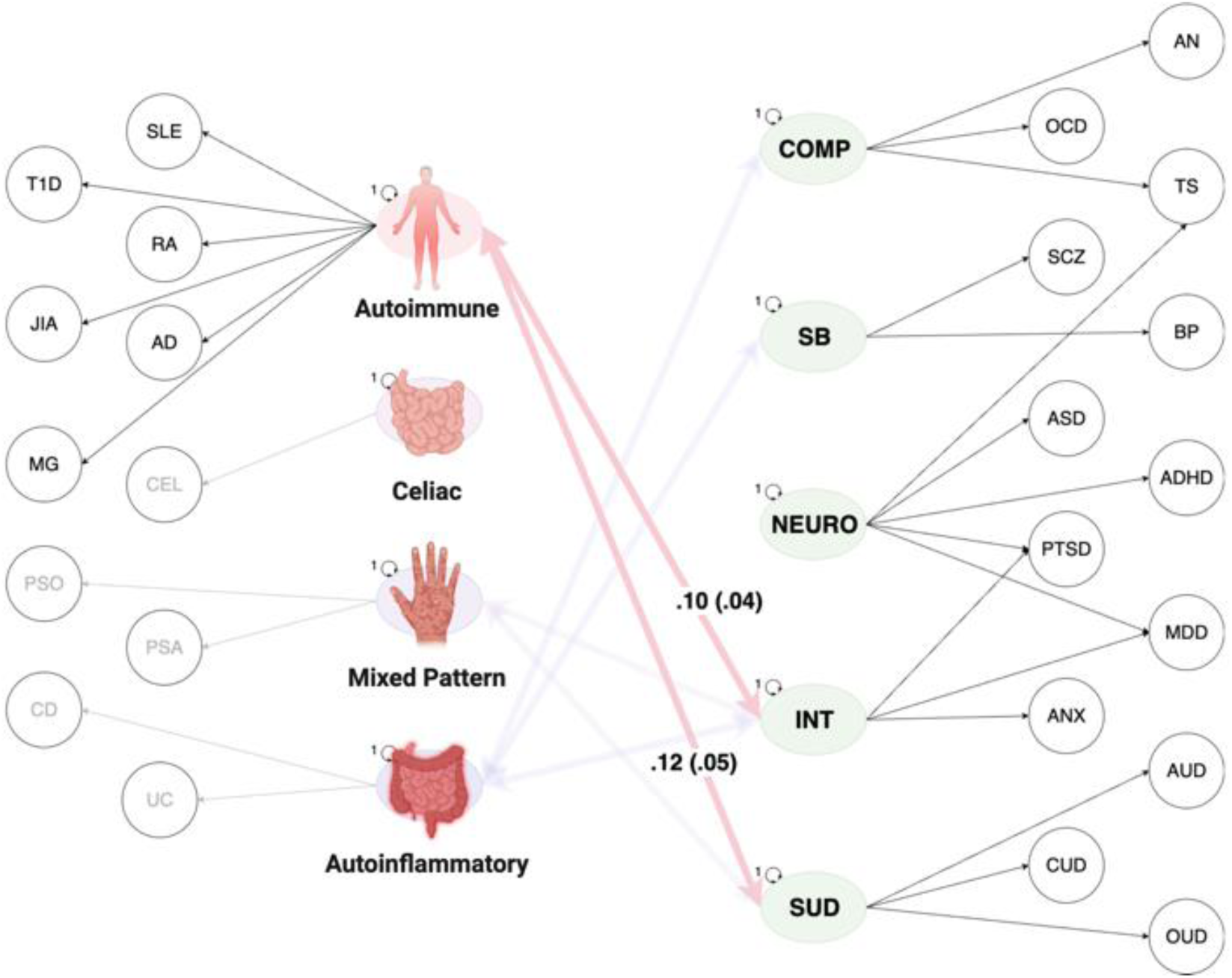
Autoimmune Diseases Correlate with Psychiatric Disorders. Path diagram with standardized estimates for the correlated factors model used in genomic structural equation modelling with four factors of immune-mediated disease and five factors of psychiatric disorders. Latent variables are represented as circles. The genetic component of each phenotype is represented with a circle because the genetic component is a latent variable that is not directly measured but is inferred using linkage disequilibrium score regression. Single-headed arrows are factor loadings; double-headed arrows connecting back to the same origin are variances; and double-headed arrows connecting two variables are correlations. All residual variances, factor loadings, non-significant correlations, as well as significant correlations with factors other than the autoimmune factor are left out for simplicity, but all factors are allowed to correlate in this model. Disease abbreviations are defined in Table 1.

### Mixed Pattern Diseases Correlate with Internalizing and Substance Use Disorders

As with the autoimmune disease factor, the mixed pattern disease factor showed positive overlap with the internalizing (*r_g_* = .08 [.03], *p_FDR_* = 3.46e-02) and substance use disorders factors (*r_g_* = .14 [.05], *p_FDR_* = 2.63e-02; **Figure 4**). The relationship between the mixed pattern diseases and the neurodevelopmental disorders was *Q_Factor_* significant (*p_FDR_* = 1.71e-02). Inspection of the pairwise genetic correlations and resigual genetic correlations (Figure S2 & **Figure 2**) revealed that this was due to positive genetic correlation between ADHD and psoriasis (*r_g_* = .15 [.01], *p_FDR_* = 5.93e-4) and a negative residual genetic correlation between ASD and psoriatic arthritis (*r_g_resid_*= -.20 [.07], *p_FDR_* = 1.73e-02). ASD stood apart from psychiatric disorders more generally with respect to a broader pattern of negative residual genetic overlap across immune-mediated diseases (**Figure 2**).

**Figure 4.**
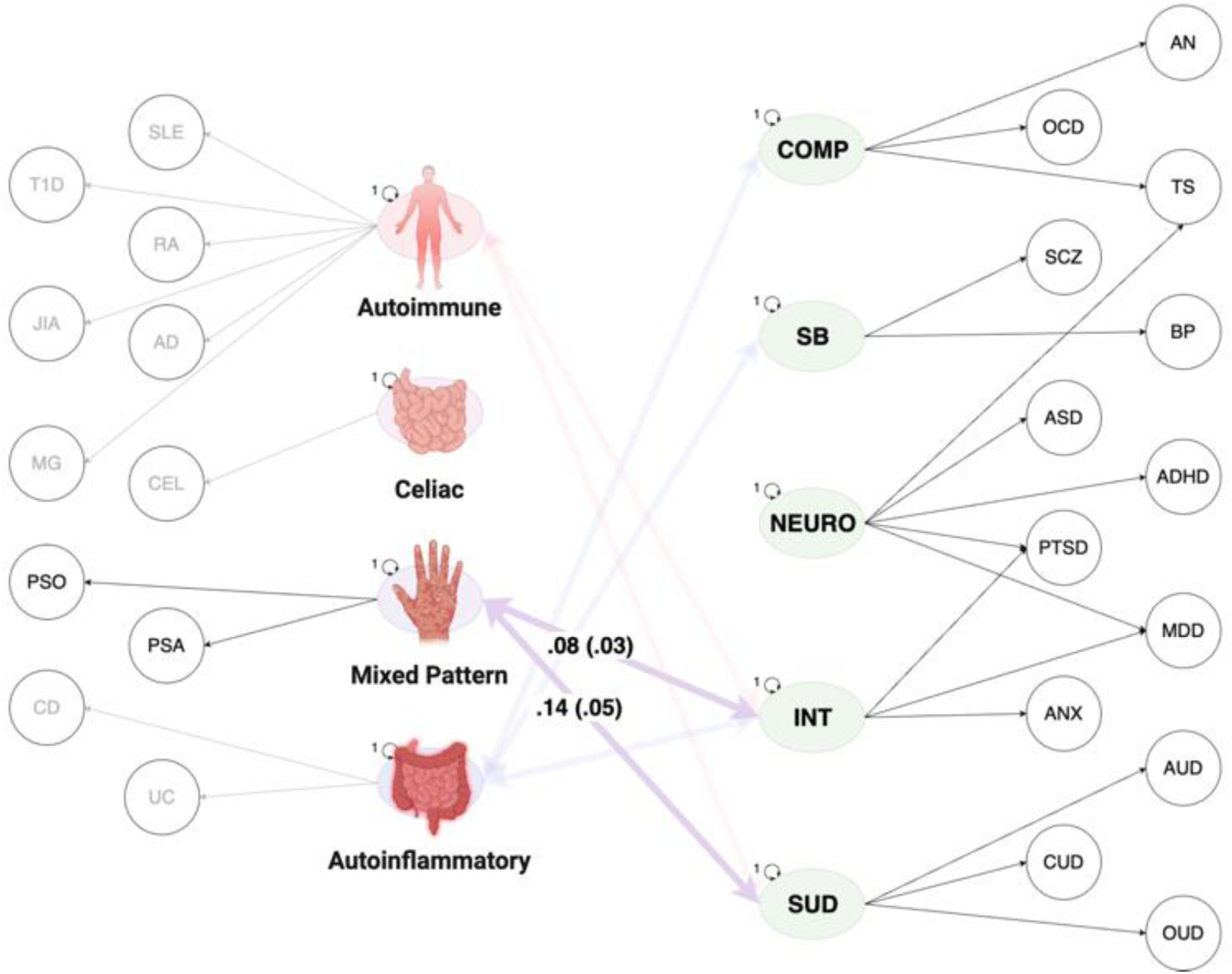
Mixed Pattern Diseases Correlate with Psychiatric Disorders. Path diagram with standardized estimates for the correlated factors model used in genomic structural equation modelling with four factors of immune-mediated disease and five factors of psychiatric disorders. Latent variables are represented as circles. The genetic component of each phenotype is represented with a circle because the genetic component is a latent variable that is not directly measured but is inferred using linkage disequilibrium score regression. Single-headed arrows are factor loadings; double-headed arrows connecting back to the same origin are variances; and double-headed arrows connecting two variables are correlations. All residual variances, factor loadings, non-significant correlations, as well as significant correlations with factors other than the mixed pattern factor are left out for simplicity, but all factors are allowed to correlate in this model. Disease abbreviations are defined in Table 1.

### Autoinflammatory Diseases Show Pervasive Overlap with Psychiatric Factors

The autoinflammatory disease factor was genetically correlated with the compulsive (*r_g_* = .13 [.06], *p_FDR_* = 3.46e-02), schizophrenia/bipolar (*r_g_*= .17 [.04], *p_FDR_* = 6.99e-05), and internalizing disorders factors (*r_g_* = .16 [.03], *p_FDR_* = 7.10e-05; **Figure 5**). No psychiatric factor correlations were *Q_Factor_* significant. Two residual genetic correlations were added between psychiatric and autoinflammatory disease for Crohn’s disease and ADHD (*r_g_* = .11 [.04], *p_FDR_* = 7.11e-03) and between ulcerative colitis and cannabis use disorder (*r_g_* = -.13 [.05], *p_FDR_*= 1.73e-02).

**Figure 5.**
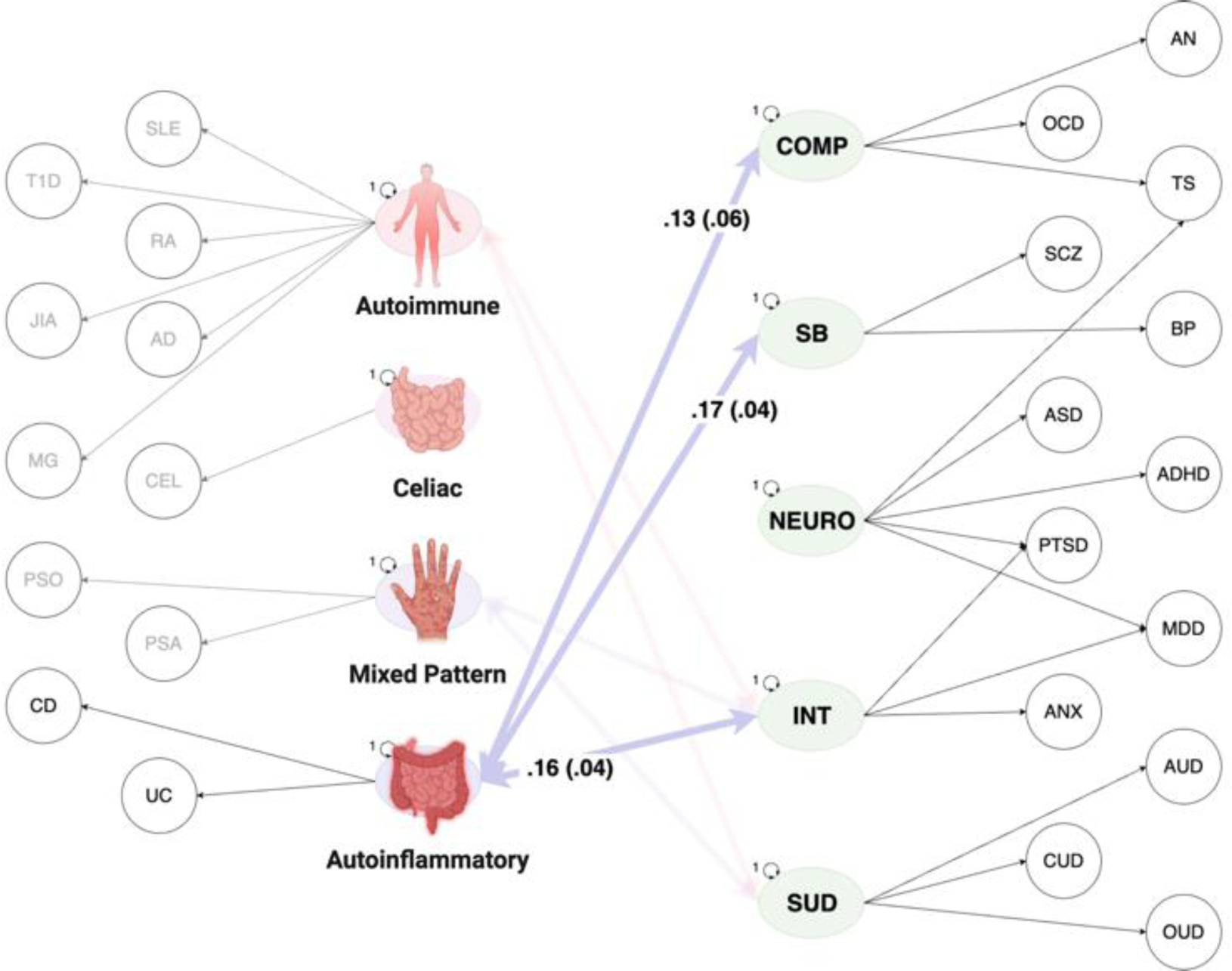
Autoinflammatory Diseases Correlate with Psychiatric Disorders. Path diagram with standardized estimates for the correlated factors model used in genomic structural equation modelling with four factors of immune-mediated disease and five factors of psychiatric disorders. Latent variables are represented as circles. The genetic component of each phenotype is represented with a circle because the genetic component is a latent variable that is not directly measured but is inferred using linkage disequilibrium score regression. Single-headed arrows are factor loadings; double-headed arrows connecting back to the same origin are variances; and double-headed arrows connecting two variables are correlations. All residual variances, factor loadings, non-significant correlations, as well as significant correlations with factors other than the autoinflammatory factor are left out for simplicity, but all factors are allowed to correlate in this model. Disease abbreviations are defined in Table 1.

## Discussion

Extant literature at phenotypic and genetic levels has established a strong link between immune-mediated disease and psychiatric disorders. Here, we applied Genomic SEM to perform the first multivariate genomic examination of immune-mediated and psychiatric disorders. We began by establishing a factor structure for immune-mediated diseases. Although the genetic factor structure of immune-mediated diseases has previously been investigated,^20,21^ we build upon these findings by incorporating additional immune-mediated diseases. Moreover, this updated factor model captures the theoretical continuum of autoimmune and autoinflammatory diseases, which allowed us to examine differential genetic overlap between clusters of psychiatric disorders with diseases characterized by the adaptive versus innate immune system. We find that a factor defined by internalizing disorders is pervasively associated with three immune-mediated factors defined by autoimmune, autoinflammatory, and mixed-pattern diseases. This is as compared to a substance use disorders factor that was specifically associated with the autoimmune factor, and compulsive and schizophrenia/bipolar factors that were only associated with the autoinflammatory factor. Additionally, we identify four significant *Q_Factor_* results that indexed psychiatric-immune relationships that did not operate via the factors, along with 14 significant residual genetic correlations between pairs of psychiatric-immune disorders. We consider these different levels of risk sharing, from broad associations across clusters of diseases to more specific risk sharing across pairs of psychiatric and immune diseases, in more detail below.

Inter-factor genetic correlations capture overlapping signal across clusters of psychiatric and immune-mediated diseases recharacterize prior bivariate findings^22^ as indexing broader patterns of risk-sharing. For example, phenotypic^23^ and genetic studies^24^ have linked depression to a host of immune-mediated diseases. Underscoring the importance of applying nuanced statistical techniques able to discern between broad and specific pathways, we find that these pairwise associations are likely indexing upstream risk pathways shared across immune-mediated diseases and other internalizing disorders. Similarly, we find that previously described genetic associations between schizophrenia^25,26^ and bipolar disorder^27^ and ulcerative colitis and Crohn’s disease reflects more extensive risk sharing across schizophrenia/bipolar and autoinflammatory factors. Etiological models of schizophrenia often include inflammatory elements and innate immunity^28^, with a recent bibliometric analysis identifying over 3,000 articles examining the link between schizophrenia and inflammation^29^. Given the sheer volume of research being conducted in this area, our findings provide a critical context for interpreting these autoinflammatory and innate risk pathways as operating at the level of what is shared across schizophrenia and bipolar disorder.

Significant residual genetic correlations between specific pairs of psychiatric and immune-mediated diseases pointed towards more circumscribed relationships across psychiatric and immune disorders. In addition, these residual genetic correlations revealed associations that were not apparent in the zero-order genetic correlations. For example, it was only when parsing out shared signal across bipolar and schizophrenia that divergent autoimmune associations emerged, with schizophrenia and bipolar showing positive (risk conferring) and negative (protective) residual associations, respectively. In particular, schizophrenia showed a high level of residual overlap with Addison’s disease. In combination with the factor level results highlighted above, autoinflammatory conditions then appear to index a component of what is shared across schizophrenia and bipolar disorder, while autoimmune conditions capture a source of genetic divergence.

Out of four total *Q_Factor_* results in our model, three were identified for the neurodevelopmental disorders factor alone, indicating that this specific psychiatric factor does not index shared pathways with immune-mediated disease. This result was driven by largely positive associations for ADHD relative to ASD’s unique pattern of negative associations with immune-mediated diseases. This finding is inconsistent with prior results linking ASD to increase risk for immune-mediated conditions^30^, though review articles have noted inconsistencies in this association^31^ or highlighted that these links may stem from the prenatal environment (e.g., maternal infection)^32^.

### Limitations and Future Directions

Due to differences in the correlational structure in the genome, the analyses presented here must be conducted within a particular genetic ancestry. GWAS data was of sufficient sample size only in individuals of European ancestry, but future work extending these analyses to other ancestral groups is of vital importance. In addition, certain autoimmune diseases, but not autoinflammatory diseases, are more predominant in females^33,34^, indicating that future analyses would also benefit from considering sex-specific genetic links between autoimmune disease and psychiatric disorders. Finally, given known biological systems and immune markers (e.g., C-reactive protein^35^) associated with immune-mediated disease, future work should seek to gain a more complete, mechanistic understanding of this psychiatric-immune link by examining the mediating role of these intermediate biological processes.

## Conclusion

The phenotypic literature describes widespread associations across pairs of immune-mediated and psychiatric disorders. The current study leveraged the unique ability of genomics to bring together different participant samples in the same statistical model to perform the first multivariate examination of these two illness domains. In doing so, our findings revealed different levels of risk sharing, from pervasive associations between immune-mediated diseases identified for internalizing disorders, to more specific relationships between certain psychiatric clusters and either the innate or adaptive immune system. Additionally, we observed genetic signal specific to individual immune-mediated diseases and psychiatric disorders, as highlighted by significant residual genetic correlations. Collectively, these results offer a more nuanced and comprehensive understanding of the different levels of risk sharing across immune-mediated diseases and psychiatric disorders.

## Data Availability

All data used in this study came from publically available sources. Summary statistics for data from the Psychiatric Genomics Consortium (PGC) can be downloaded or requested here: https://www.med.unc.edu/pgc/download-results/;
Summary statistics for the Anxiety phenotype can be downloaded here: https://drive.google.com/drive/folders/1fguHvz7l2G45sbMI9h_veQun4aXNTy1v;
Summary statistics for Celiac Disease: http://ftp.ebi.ac.uk/pub/databases/gwas/summary_statistics/GCST90014001-GCST90015000/GCST90014442/;
Summary statistics for Systemic Lupus Erythematosus: http://ftp.ebi.ac.uk/pub/databases/gwas/summary_statistics/GCST003001-GCST004000/GCST003156/;
Summary statistics for Diabetes Type 1: http://ftp.ebi.ac.uk/pub/databases/gwas/summary_statistics/GCST90014001-GCST90015000/GCST90014023/;
Summary statistics for Rheumatoid Arthritis: http://plaza.umin.ac.jp/~yokada/datasource/software.htm;
Summary statistics for Crohn's Disease: http://ftp.ebi.ac.uk/pub/databases/gwas/summary_statistics/GCST004001-GCST005000/GCST004132/;
Summary statistics for Ulcerative colitis: http://ftp.ebi.ac.uk/pub/databases/gwas/summary_statistics/GCST004001-GCST005000/GCST004133/;
Summary statistics for Juvenile idiopathic arthritis: http://ftp.ebi.ac.uk/pub/databases/gwas/summary_statistics/GCST90010001-GCST90011000/GCST90010715/harmonised/;
Summary statistics for Addison's Disease: http://ftp.ebi.ac.uk/pub/databases/gwas/summary_statistics/GCST90011001-GCST90012000/GCST90011871/harmonised/;
Summary statistics for Myasthenia gravis: http://ftp.ebi.ac.uk/pub/databases/gwas/summary_statistics/GCST90093001-GCST90094000/GCST90093061/;
Summary statistics for Psoriasis: http://ftp.ebi.ac.uk/pub/databases/gwas/summary_statistics/GCST90019001-GCST90020000/GCST90019016/;
Summary statistics for Psoriatic arthritis: http://ftp.ebi.ac.uk/pub/databases/gwas/summary_statistics/GCST90243001-GCST90244000/GCST90243956/;
Links to the LD-scores, reference panel data, and the code used to produce the current results can all be found at: https://github.com/GenomicSEM/GenomicSEM/wiki

## Acknowledgements

SB, ADG, JML, LSS are supported by NIMH Grant R01MH120219. ADG is supported by NIA Grant RF1AG073593. SB is supported by the Shurl and Kay Curci Foundation.

