## Supplementary material for "Examining the Genetic Links between Clusters of Immune-mediated Diseases and Psychiatric Disorders": Online Supplement

***Q_Factor_ Heterogeneity Test***

*Overview.* In the context of the current paper we introduce and validate *Q_Factor_*, a heterogeneity index that serves to identify sources of local misfit between two latent genomic factors. More specifically, by utilizing *Q_Factor_* we can identify factors that appear to be significantly genetically correlated on an overall factor level, but these factor correlations are specifically driven by a subset of indicators (traits) that load on either factors. Conversely, *Q_Factor_* can also identify factors that are not significantly genetically correlated because the indicators across factors are, on average, not genetically correlated, but are still defined by indicators with substantial genetic overlap across factors. More broadly, *Q_Factor_* helps detect relationships between cross-trait, cross-factor relationships that are more circumscribed and do not operate at the level of the genetic risk sharing captured by the factors they define.

*Q_Factor_* is calculated by taking the residual covariance matrix for the subset of the matrix that reflects the cross-factor associations for the traits that define the two correlated factors (*R_Factor_*) as:

*R*_Factor_ = $S_{Factor}- \sum\theta_{Factor}$	(1)

where $S_{Factor}$is a subset of the full LDSC estimated genetic covariance matrix (*S*) and $\sum\theta_{Factor}$ a subset of the full model implied covariance matrix ($\sum\theta).$ In both instances, they are subset to only the cross-factor associations at the level of the traits that define the factors. The model implied estimates, $\sum\theta_{Factor,}$ for each pair of traits reflects the product of the factor loading for one trait on the first factor, the factor correlation, and the factor loading for a separate trait on the second factor.

To further illustrate this point, an exemplar $S_{Factor}$is depicted below for a factor model where one factor is defined by the genetic signal for three indicators (*g1, g2, g3*) and a second, correlated factor is defined by the genetic signal for three separate indicators (*g4, g5, g6*). While the full 6$\times6$ genetic covariance matrix (*S*) would include the 6 SNP-based heritabilities on the diagonal and the 15 genetic covariances on the off-diagonal, for the purposes of calculating *Q_Factor_* this full genetic covariance matrix is subset to the lower block reflecting the 9 genetic covariances across traits that load on separate factors:

$S_{Factor} =\left[ \begin{matrix} & & & & & \\ - & & & & & \\ - & - & & & & \\ - & - & - & & & \\ \sigma_{g1,g4} & \sigma_{g2,g4} & \sigma_{g3,g4} & - & & \\ \sigma_{g1,g5} & \sigma_{g2,g5} & \sigma_{g3,g5} & - & - & \\ \sigma_{g1,g6} & \sigma_{g2,g6} & \sigma_{g3,g6} & - & - & - \end{matrix} \right]$	(2)

Taken together, *R*_Factor_ then represents the extent to which a single factor correlation is insufficient for describing the patterning of relationships between the traits across factors. The precision of these cross-factor relationships is indexed by taking the eigendecomposition of the portion of the sampling covariance matrix (*V*) that indexes those cross-factor associations (*V_Factor_*):

*V_Factor_* = (P_1_ P_0_) $\binom{E 0}{0 0}$ $\left( \begin{aligned} P_{1}^{'} \\ P_{o}^{'} \end{aligned} \right)$	(3)

where *P_1_* is the matrix of principal components (eigenvectors) of *V_Factor_*, *P_0_* is the null space of *V_Factor_*, and $E$ is a diagonal matrix of the non-zero eigenvalues of *V_Factor_*. These eigenvalues and eigenvectors can then be used to weight the residual covariance matrix of the cross-factor, cross-trait estimates to obtain a $\chi$^2^ distributed test statistic given as:

*Q_Factor_*$\left( df \right)\sim R_{Factor}'$P_1_$E$^-1^P_1_${'R}_{Factor}$	(4)

where *df* reflects the degrees of freedom, which will be one less than the number of cross-factor, cross-trait associations (e.g., for the equation 2 above, the *df* would be 8). We note that the open-source Genomic SEM R package will iteratively apply this equation for each estimated inter-factor correlation in the model, and in doing so will calculate separate *R*_Factor,_ *V_Factor,_* and *Q_Factor_* estimates.

*Simulations.* We ran a series of simulations to validate the statistical properties of *Q_Factor ._* Across all simulations, the population generating model consisted of two, six-item factors. For each factor, one trait had a standardized factor loading of .15, .2, .3, .4, .5, or .7. We then ran two sets of 1,000 simulations (2,000 simulations total) consisting of 50,000 participants for each simulation. These sets of simulations differed only with respect to whether the generating population specified the two factors to correlated at either .2 or .4. As the cross-trait, cross-factor relationships were specified to operate entirely through the factor, these reflect a null generating population for *Q_Factor_,* which allowed us to verify that it conformed to a $\chi^{2}$ distributed test statistic and that it had a well-calibrated Type I error (false positive) rate. We highlight that for a $\chi^{2}$ test the null is equal to the *df,* where the *df* for this particular example is 35. The mean *Q_Factor_* estimate across the simulations with a factor correlation of .2 was 33.77 [*SD* = 8.18] with a well-controlled Type I error rate of 3.9% of runs producing an estimate that was significant at *p* < .05. The mean *Q_Factor_* across simulations for a factor correlation of .4 was 31.55 [*SD* = 7.61] with a well-controlled Type I error rate of 1.8% of runs significant at *p* < .05. The distributions of *Q_Factor_* estimates across the 1,000 simulations for factor correlations of .2 and .4 are given in Figures S3 and S4 respectively and can be seen to closely conform to a $\chi^{2}$distribution. Taken together, these results indicate that *Q_Factor_* is well-calibrated test statistic for evaluating the ability of a single factor correlation to describe the cross-trait, cross-factor associations.

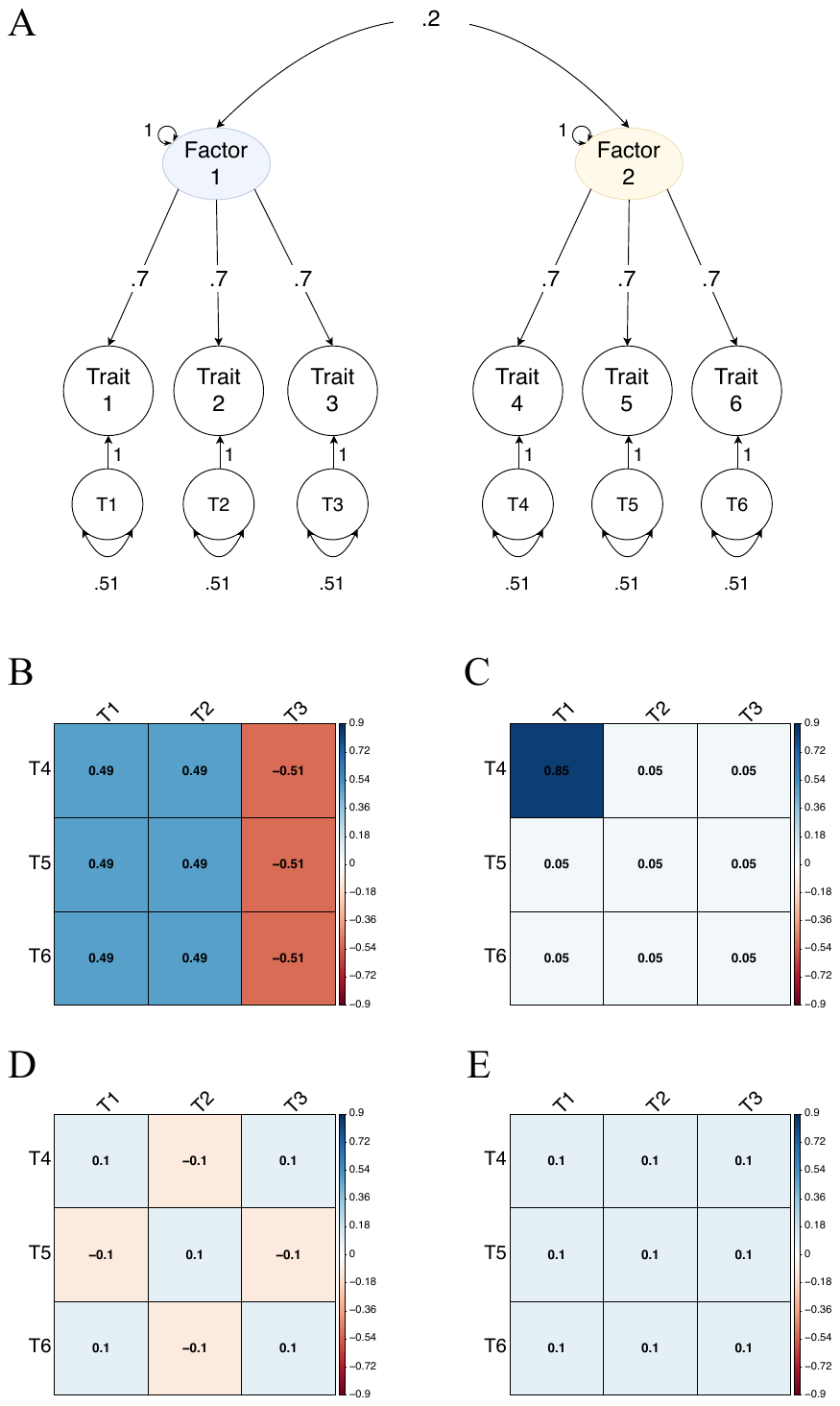
**Supplementary Figure 1. Hypothetical *Q_Factor_* Illustration**. *Panel A* depicts a path diagram of two theoretical factors, factor 1 and factor 2. Trait 1-3 load onto factor 1 and trait 4-6 load onto factor 2, with the two factors correlating at .2. Panels B-E depict the observed genetic correlation matrices between the factor 1 and factor 2 traits, wherein the pattern of correlations within each of these matrices implies the depicted factor correlation of .2. Panel B-D reflect different scenarios where Q_Factor_ would be significant as a single factor correlation is insufficient for describing the pattern of relationships across the factor 1 and factor 2 traits. *Panel B.* Trait 3 on factor 1 is negatively genetically correlated with all traits on factor 2, while traits 1 and 2 are positively correlated with traits on factor 2. These divergent associations are misrepresented by a single factor correlation of .2, which is why Q_Factor_ is significant. *Panel C.* Trait 1 on factor 1 and trait 4 on factor 2 are very strongly correlated, while all other traits on the two factors are weakly genetically correlated, thus the overall factor correlation is driven by one trait on each factor and this factor correlation is consequently identified as Q_Factor_ significant. *Panel D*. There is overall divergence within the correlation matrix in how different traits on factor 1 are correlated with different traits on factor 2, which produces a significant Q_Factor_ metric. *Panel E*. This is the one depicted scenario where Q_Factor_ would not be significant as the pattern of relationships across the factor 1 and factor 2 traits, which consists of uniformly positive correlations of an equal size that is consistent with the equally sized factor loadings, is appropriately represented by a single factor correlation. We note that when the factor loadings are different across traits that a single factor correlation can still be consistent with differently sized correlations across traits so long as the divergences across correlations scale with the divergences across loadings.

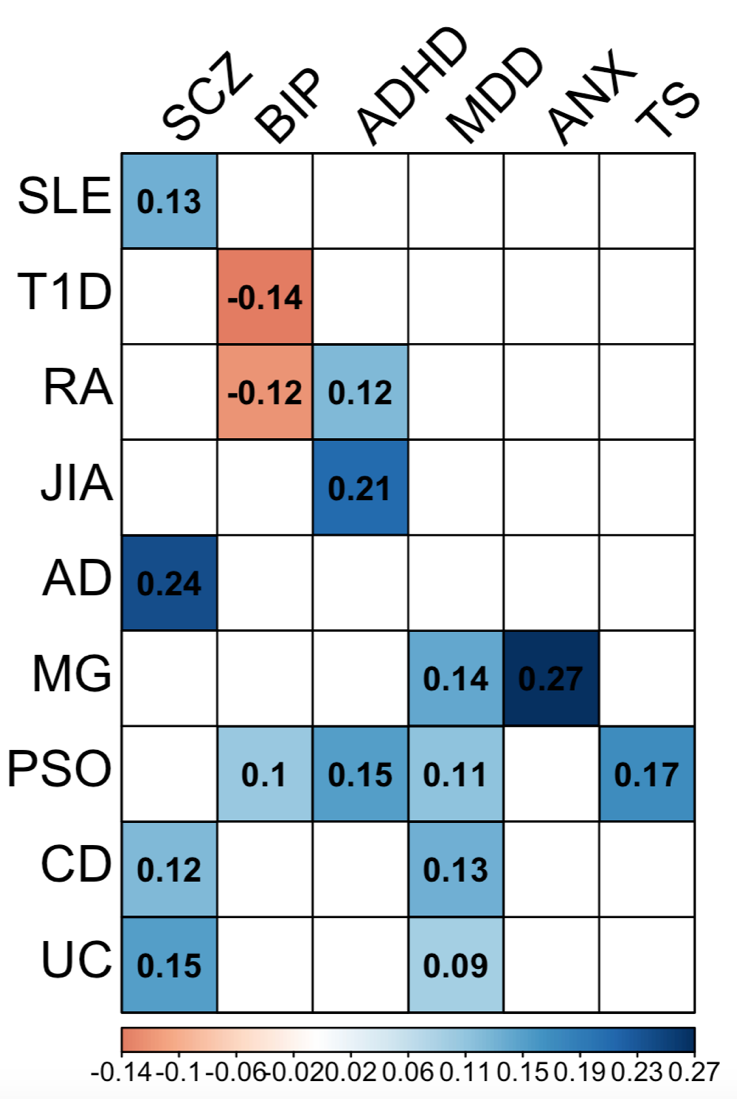

**Supplementary Figure 2. Significant Genetic Correlations**. Depicts significant genetic correlations in the original correlation matrix. SLE, systemic lupus erythematosus; T1D, diabetes type 1; RA, rheumatoid arthritis; JIA, juvenile idiopathic arthritis; AD, Addison’s disease; MG, myasthenia gravis; CEL, celiac disease; PSO, psoriasis; PSA, psoriatic arthritis; UC, ulcerative colitis; CD, Crohn's disease; ADHD, attention-deficit/hyperactivity disorder; ANX, anxiety disorders; ASD, autism spectrum disorder; BIP, bipolar disorder; CUD, cannabis use disorder; MDD, major depressive disorder; OCD, obsessive-compulsive disorder; PTSD, posttraumatic stress disorder; TS, Tourette’s syndrome.

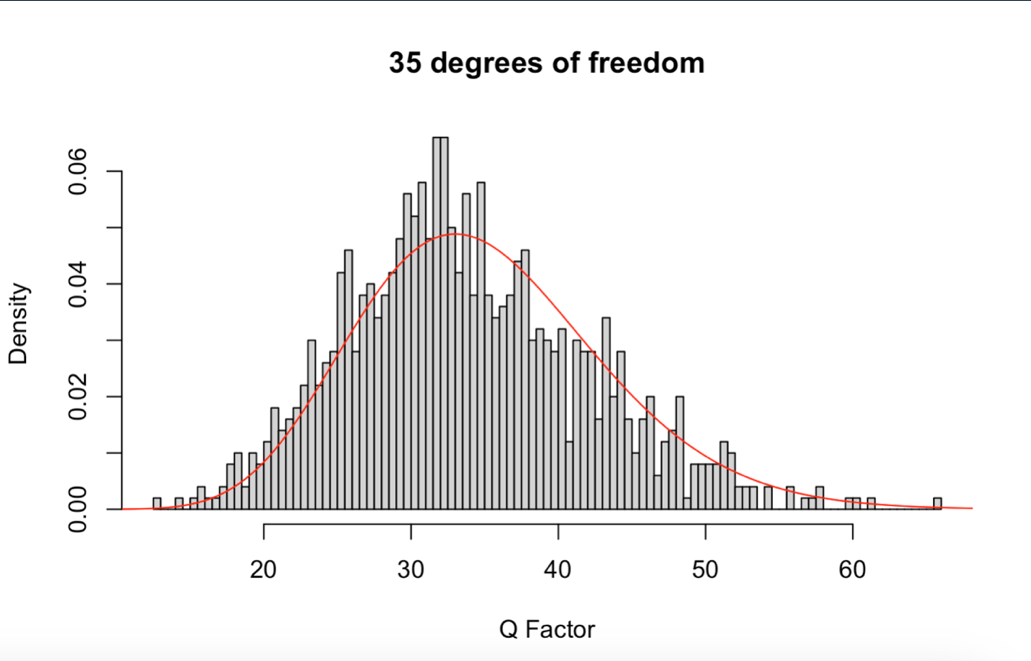

**Supplementary Figure 3. Q_Factor_ Simulation for Factor Correlation of .2.** The bars in the histogram depict the different Q_Factor_ estimates for 1,000 simulations when the generating population had a factor correlation of 0.2. The expected $\chi^{2}$ distribution is given in red.

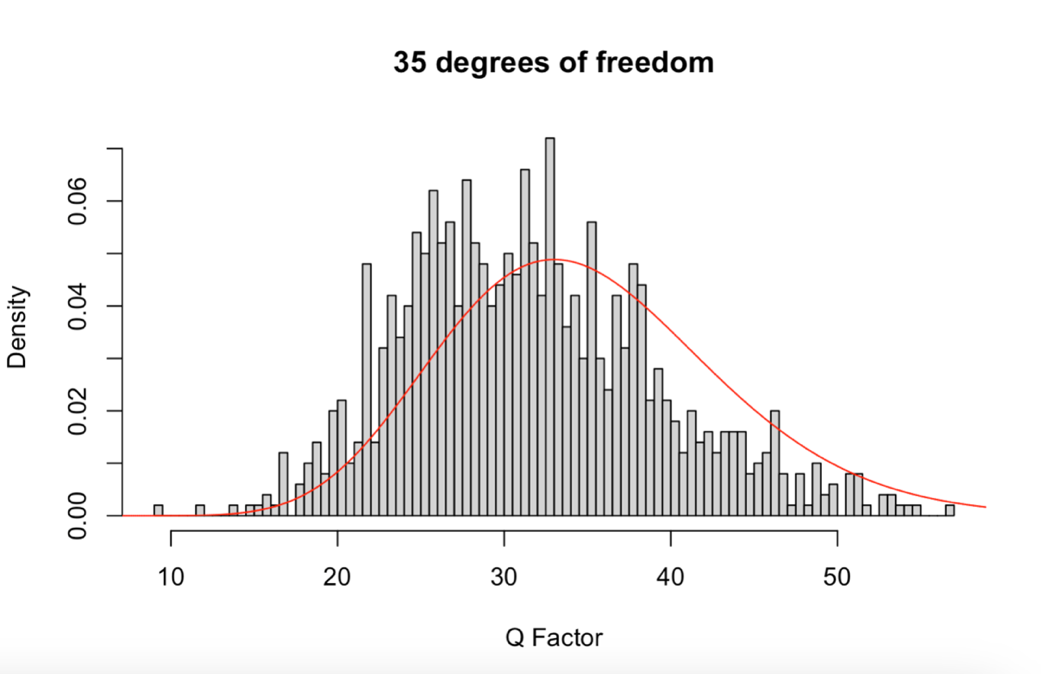

**Supplementary Figure 4. Q_Factor_ Simulation for Factor Correlation of .4.** The bars in the histogram depict the different Q_Factor_ estimates for 1,000 simulations when the generating population had a factor correlation of 0.4. The expected $\chi^{2}$ distribution is given in red.
